# A Novel 3-Dimensional technique in measuring pericoronary epicardial adipose tissue radiodensity

**DOI:** 10.1101/2020.10.29.20222679

**Authors:** Lingyu Xu, Stanislau Hrybouski, Yuancheng Xu, Richard Coulden, Emer Sonnex, D. Ian Paterson, Craig Butler

## Abstract

**Objectives:** This study aimed to investigate a novel semi-automated three-dimensional (3D) quantification of the pericoronary epicardial adipose tissue radiodensity (PCATrd).

**Methods:** Twenty-four subjects who previously underwent contrast-enhanced cardiac CT scans were retrospectively identified. The PCATrd was measured in ITK-SNAP imaging software using a Hounsfield unit threshold (−190,-3) to define epicardial adipose tissue (EAT). A spherical 3D brush tool was used on multiplanar reformatted images to segment the PCAT. We defined the PCATrd as EAT within the orthogonal distance from the coronary artery (CA) outer wall equal to the diameter of the corresponding CA segment. The segmentation followed the path of major CAs. Additionally, the PCAT of twenty-five calcified segments were segmented. Reliability of this novel segmentation protocol was assessed using Dice Similarity Coefficients (DSCs) and intraclass coefficient (ICC).

**Results:** The segmentation reproducibility for the PCAT was high, with intraobserver DSC 0.86±0.04 for the full length of major CAs and 0.85±0.07 for the calcified segments, and interobserver DSC 0.84±0.04 for the full length of major CAs and 0.83±0.05 for the calcified segments. The reproducibility of the PCATrd value assessed by ICC was also excellent, with intraobserver ICC 0.99 for the full length of major CAs and 0.99 for the calcified segments, and interobserver ICC 0.99 for the full length of major CAs and 0.99 for the calcified segments.

**Conclusions:** Our novel 3D PCATrd quantification technique is reliable and reproducible. The availability of the open source software and detailed image analysis pipeline will enable reliable replications and broad uptake of our technique.

**Key points:**

- We have produced a novel, semiautomated technique to comprehensively quantify pericoronary epicardial adipose tissue radiodensity (PCATrd) which is a novel imaging biomarker of coronary inflammation.
- Our method of PCAT segmentation has excellent reproducibility.
- We use open source software and provide detailed image analysis pipeline of quantifying PCATrd, which will allow easy replication and broad uptake of our technique.

## 1. Introduction

Epicardial adipose tissue (EAT) can be used to assess cardiovascular risk through its influence on the development of coronary atherosclerosis via paracrine secretion of pro-atherosclerotic cytokines [1]. Methods for characterizing EAT have changed over recent years and have moved away from volume-based assessments [2; 3] to tissue characterization such as radiodensity, and more recently, pericoronary EAT radiodensity (PCATrd). EAT radiodensity is associated with cardiovascular disease and adverse outcomes [3-5]. EAT radiodensity shows heterogeneity throughout the heart [6-8], undermining the representativeness of current techniques that use regional sampling [7] for estimating the EAT radiodensity. Current evidence suggests that the PCATrd reflects differences in local inflammatory conditions, which exhibit bidirectional signalling between the pericoronary EAT and coronary arteries (CA) themselves [6]. In particular, the radiodensity of EAT closest to a CA (i.e., PCATrd) has a stronger association with cardiovascular disease and adverse outcomes than the EAT, which is distant to CA[6; 7]. The variability in PCATrd is a key reason why we postulate that developing a technique for a comprehensive characterization of the PCATrd over the majority of the CA tree is more informative than methods that rely on incomplete PCATrd sampling.

Antonopoulos et al. recognized the value of comprehensive PCATrd characterization but restricted their assessment to only the proximal 4 cm of the CAs and used proprietary tools [6; 7]. The generalizability of the first 4 centimeters to the entire coronary tree is unknown and runs counter to the value of a complete coronary assessment, such as coronary calcification, which robustly predicts cardiovascular risk [8; 9]. We also think that clinical studies employing a comprehensive assessment of the PCATrd will be more closely associated with CA disease than techniques using limited PCAT sampling [9; 10]. Consequently, this study’s main goal was to develop a standardized and reliable PCAT segmentation protocol, which can be used to quantify the PCATrd throughout the entire length of major epicardial CAs.

## 2. Materials and methods

### 2.1 Patient Population

We based our sample size calculations on the selection of two-sided 95% confidence interval width of the interclass correlation coefficient (ICC) for the variable General PCATrd, [11] therefore the width of confidence intervals around ICC should be no wider than 0.1,[12] therefore we calculated a range of samples sizes between 0.08 and 0.1. Typically, ICC values of over 0.9 are considered excellent[13], and our preliminary work led us to estimate our mean ICC to be 0.95. Lastly we specified a power of 0.9 and an alpha of 0.05. Our calculation produced a range of sample size from 16 (for 95% confidence interval width of 0.1) and 24 for 95% confidence interval with of 0.08. We opted for the more conservative estimate of sample size of 24.

Twenty-four subjects were randomly selected from a local research database, and none were excluded based on image quality [14]. The inclusion criterion included 1) coronary computed tomography angiography (CCTA) scans imaged at the same tube energy (i.e., 120 KV in current study); 2) a minimum scan coverage extending from the pulmonary artery bifurcation to ventricular apex in the z-axis; 3) and imaging datasets were reconstructed with standard parameters (i.e. same slice thickness and interslice gap). Ethical approval was obtained from our institution’s human research ethics committee, and written consent form was obtained from each participant.

### 2.2 MDCT protocol

All subjects underwent contrast-enhanced cardiac CT scans in a multidetector CT scanner (Somatom Sensation 64, Siemens, Germany) using a standardized acquisition protocol as previously described [14; 15]. During the CT scan, the participants’ heart rate was controlled at 70 bpm or less following pretreatment with oral metoprolol, as necessary. The CT scan was triggered prospectively at 70% of the RR interval or retrospectively with a 120-kV tube voltage. CTA was performed following intravenous injection of 75ml (rate: 6 ml/s; iodine delivery rate: 2.1 g I/s) of iodine-based contrast (Omnipaque 350, GE Healthcare, USA). Reconstructed images consisted of 0.75-mm thick slices with a 0.4-mm interslice gap.

### 2.3 Assessment of pericoronary epicardial adipose tissue radiodensity (PCATrd)

The details of the PCATrd analysis pipeline was described in the Supplemental material 1. Briefly, we described the pipeline analysis as the following.

Step 1: The CT dataset was uploaded to the dedicated image analysis software (ITK-SNAP, Version 3.8.0) [16]; the targeted CCTA image series were selected and saved as NIFTI file;.

Step 2: Create the Hounsfield Unit defined threshold map of EAT in MATLAB software (MATLAB R2017b), using our in-house MATLAB code (See Supplemental material 1). The radiodensity threshold was derived from our previously study[15], ranging from -190 to -3 HU to define EAT in contrast cardiac CT datasets.

Step 3: Apply this threshold map to the CCTA images in ITK-SNAP software.

Step 4: Start the segmentation of PCATrd.

Briefly, CCTA images with threshold map were three-dimensionally displayed (axial, coronal, and sagittal planes) in ITK-SNAP software. The PCAT was defined as the EAT immediately adjacent to the CAs, specifically a three-dimensional cylindrical layer with a radial distance from the outer coronary vascular wall equal to the diameter [6; 7] of the corresponding coronary segment. A spherical 3D brush tool was used to manually segment the PCAT in three orthogonal planes simultaneously (Figure 1A). Tracing of PCAT was propagated down the length of the coronary artery by using the 3D brush tool with position referencing in all three orthogonal planes and with 2-3 mm intervals between each consecutive tracing. The PCAT was segmented along the major length of the left main (LM), the left anterior descending (LAD), left circumflex (LCX) arteries and the right CA (RCA). A 3D visualization of segmented PCAT from one of our participants is shown in Fig. 1D. PCATrd was computed by averaging radiodensity values across all voxels belonging to a given PCAT label. Finally, we computed general PCATrd by collapsing all 4 PCAT segments (i.e., LM, LAD, LCX, and RCA) into a single label.

**Figure 1.**
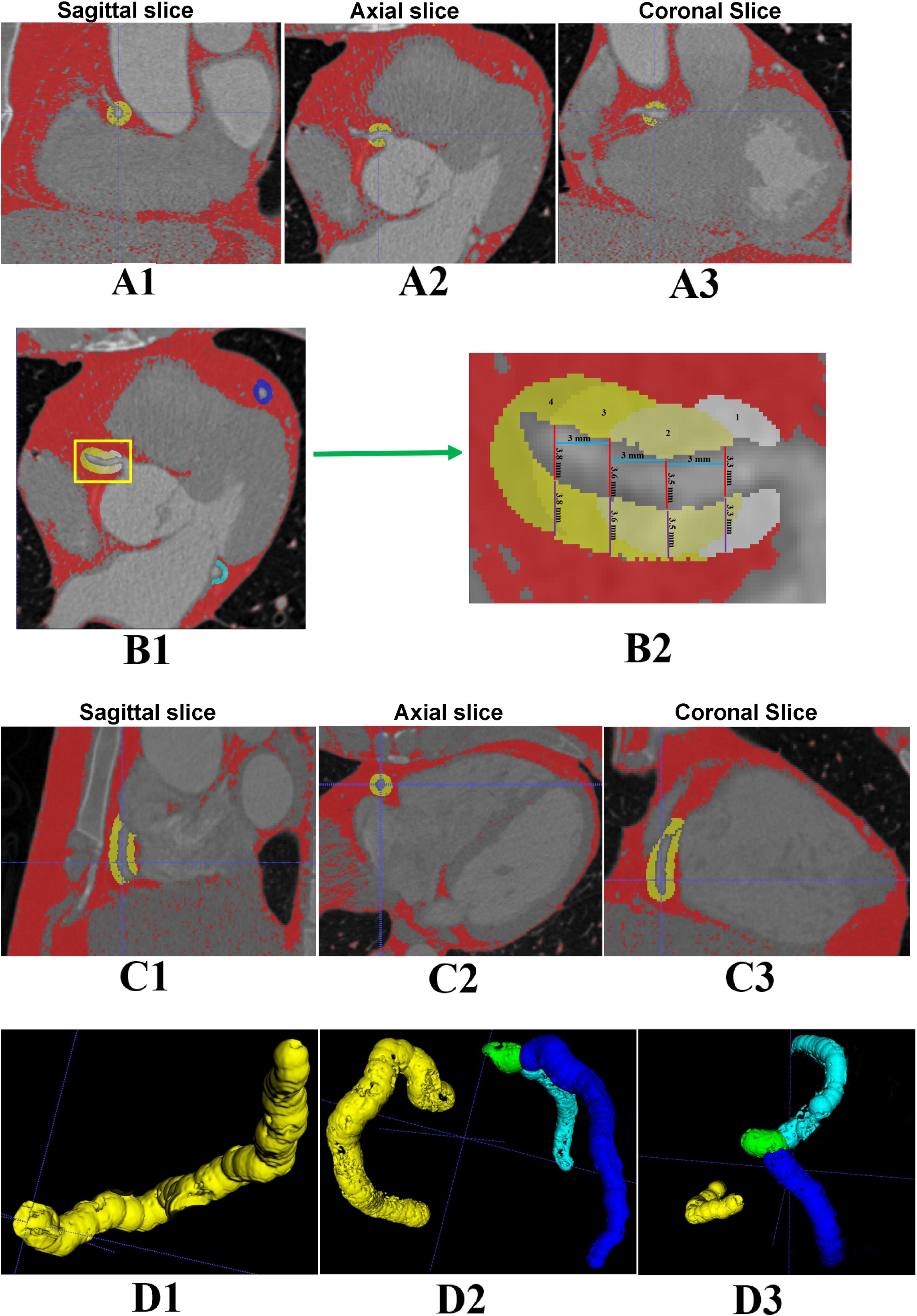
Description of 3D PCAT segmentation. A1-A3: The 3D cursor was placed at the center of RCA in 3 orthogonal images simultaneously. B1: Advance the 3D cursor with an interval of 3 mm (light blue line) by following the CA; Make the segmenting radius of PCAT (Purple line) equal to the diameter of corresponding CA (Red line). B2: The yellowish square part in B1 was amplified. C1-C3: Segment the full RCA PCAT from the ostium to the origin of the posterior descending artery at 3 orthogonal images, respectively. D1-D3: 3D renderings of completed RCA (yellow), LM (green), LAD (blue), and LCX (cyan) PCAT tracings. D1 shows RCA only; D2 and D3 are reconstructions of all CA segments viewed from left to right, and superior to inferior directions, respectively. Abbreviations: see Table 1

### 2.4 Assessment of pericoronary epicardial adipose tissue radiodensity (PCATrd) of the calcified segments

Using the same technique outlines in section 2.3, we performed an additional PCATrd assessment in 25 calcified coronary segments to assess the effect of coronary calcification/plaque on the reliability of PCATrd estimates. We segmented the PCAT along the length of calcified segments with additional 1 mm at both the proximal and the distal end.

**Table 1.**
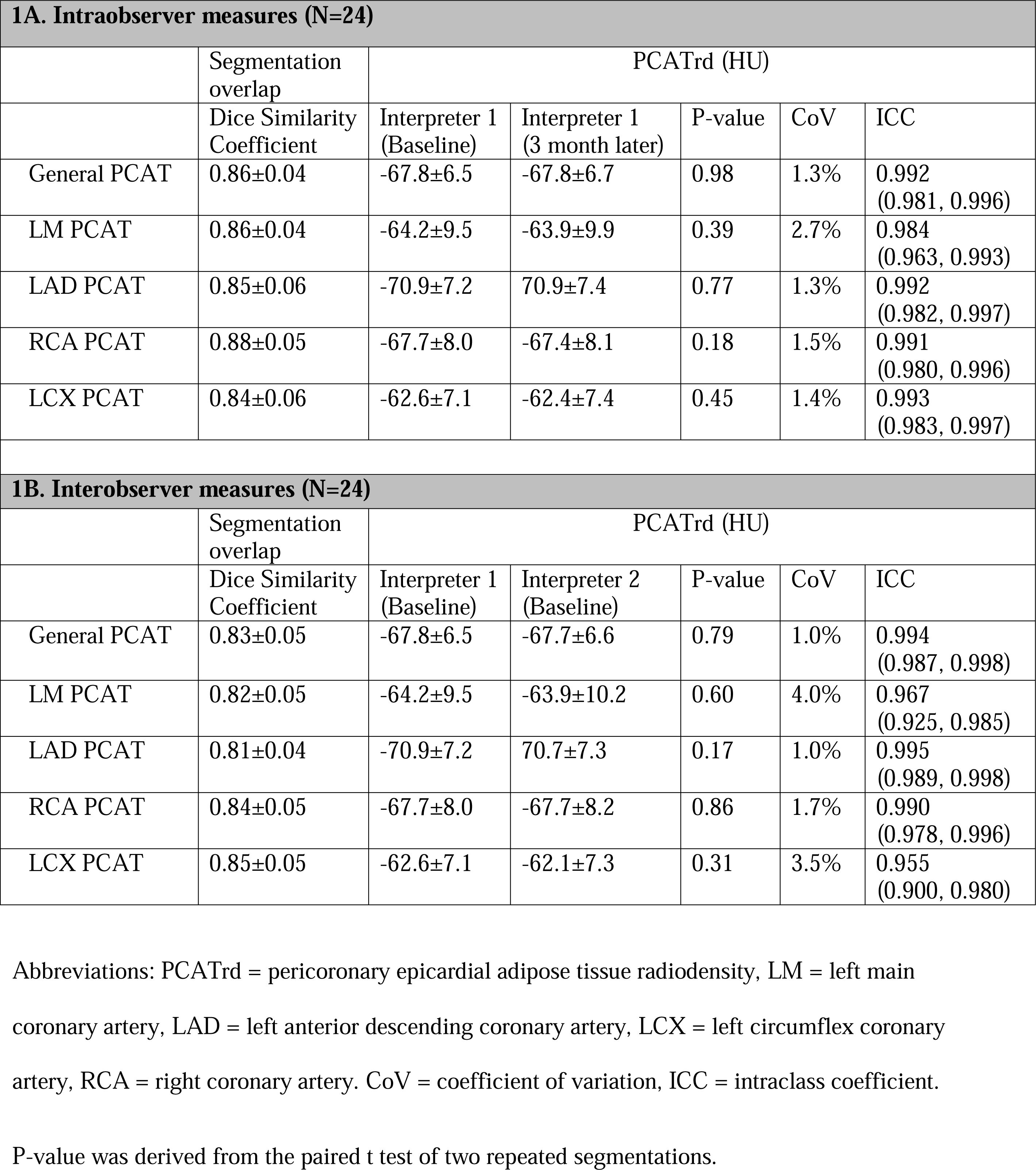
Intraobserver and interobserver measures of PCAT.

| **1A. Intraobserver measures (N=24)** |  |  |  |  |  |  |
| --- | --- | --- | --- | --- | --- | --- |
|  | Segmentation overlap | PCATrd (HU) |  |  |  |  |
|  | Dice Similarity Coefficient | Interpreter 1 (Baseline) | Interpreter 1 (3 month later) | P-value | CoV | ICC |
| General PCAT | 0.86±0.04 | -67.8±6.5 | -67.8±6.7 | 0.98 | 1.3% | 0.992<br>(0.981, 0.996) |
| LM PCAT | 0.86±0.04 | -64.2±9.5 | -63.9±9.9 | 0.39 | 2.7% | 0.984<br>(0.963, 0.993) |
| LAD PCAT | 0.85±0.06 | -70.9±7.2 | 70.9±7.4 | 0.77 | 1.3% | 0.992<br>(0.982, 0.997) |
| RCA PCAT | 0.88±0.05 | -67.7±8.0 | -67.4±8.1 | 0.18 | 1.5% | 0.991<br>(0.980, 0.996) |
| LCX PCAT | 0.84±0.06 | -62.6±7.1 | -62.4±7.4 | 0.45 | 1.4% | 0.993<br>(0.983, 0.997) |
| **1B. Interobserver measures (N=24)** |  |  |  |  |  |  |
|  | Segmentation overlap | PCATrd (HU) |  |  |  |  |
|  | Dice Similarity Coefficient | Interpreter 1 (Baseline) | Interpreter 2 (Baseline) | P-value | CoV | ICC |
| General PCAT | 0.83±0.05 | -67.8±6.5 | -67.7±6.6 | 0.79 | 1.0% | 0.994<br>(0.987, 0.998) |
| LM PCAT | 0.82±0.05 | -64.2±9.5 | -63.9±10.2 | 0.60 | 4.0% | 0.967<br>(0.925, 0.985) |
| LAD PCAT | 0.81±0.04 | -70.9±7.2 | 70.7±7.3 | 0.17 | 1.0% | 0.995<br>(0.989, 0.998) |
| RCA PCAT | 0.84±0.05 | -67.7±8.0 | -67.7±8.2 | 0.86 | 1.7% | 0.990<br>(0.978, 0.996) |
| LCX PCAT | 0.85±0.05 | -62.6±7.1 | -62.1±7.3 | 0.31 | 3.5% | 0.955<br>(0.900, 0.980) |

Abbreviations: PCATrd = pericoronary epicardial adipose tissue radiodensity, LM = left main coronary artery, LAD = left anterior descending coronary artery, LCX = left circumflex coronary artery, RCA = right coronary artery. CoV = coefficient of variation, ICC = intraclass coefficient.
P-value was derived from the paired t test of two repeated segmentations.

### 2.5 Reproducibility

Dice Similarity Coefficient (DSC), which can range from 0 to 1 and represents the extent of voxel overlap across different tracings, was used to assess the intraobserver and interobserver reproducibility of our PCAT segmentation protocol on all 12 subjects (i.e., 48 CAs). Custom MATLAB functions were used to calculate Dice Similarity Coefficients using equation 1:

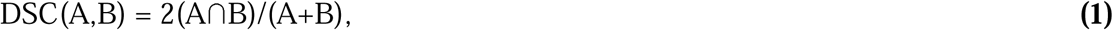

where A and B represent the number of PCAT voxels in each reliability assessment, and A∩B represents the number of voxels labeled as PCAT in both tracings [17]. Good segmentation reproducibility is obtained if DSC > 0.70, and DSC > 0.80 is indicative of excellent reliability [18]. Intra-class coefficient (ICC) with 95% confidence interval, coefficient of variation (CoV), and Bland-Altman analyses were used for assessing intraobserver and interobserver PCATrd reproducibility.

Intraobserver measurements of PCAT and PCATrd were based on two repeated measurements with a three-month interval by LX. Interobserver segmentations were performed independently by Interpreters LX and SH.

### 2.6 Statistics

The data was analyzed using commercially available statistical software (STATA version 16.0; StataCorp LP, College Station, Texas, USA). Continuous variables were expressed as mean ± standard deviation (SD) or median (25^th^, 75^th^ percentile), as appropriate. Categorical variables were expressed as frequencies and percentages. Paired t-test was used to compare PCATrd with repeated measurements. A *p*-value less than 0.05 was considered statistically significant in all tests.

## 3 Results

### 3.1 Basic characteristics

Twenty-four subjects – mean age of 55.1±8.4 years, mean body mass index of 25.4 ± 4.3 kg/m^2^, 50% males, 42% with documented coronary artery disease – passed our selection criteria and were selected for the study. The mean segmented length of PCAT was 11.5±3.5 mm in the LM, 98.7± 19.1 mm in the LAD, 73.0±10.7 mm in the RCA and 71.3±12.6 mm in the LCX. The average overall segmented length per subject was 254.5±26.3mm. The average analysis time for both interpreters was 7-9 minutes for the LAD, 5-6 minutes for each of RCA and LCX, and 1-2 minutes for the LM (i.e., 17-23 minutes per subject).

### 3.2 Intraobserver Reproducibility of PCATrd

The intraobserver segmentation overlap (Fig. 2A) was excellent, with DSC 0.85±0.04 for the overall PCAT, 0.85±0.04 for the LM PCAT, 0.84±0.07 for the LAD PCAT, 0.87±0.05 for the RCA PCAT, and 0.82±0.07 for the LCX PCAT (Fig. 2A).

**Figure 2A:**
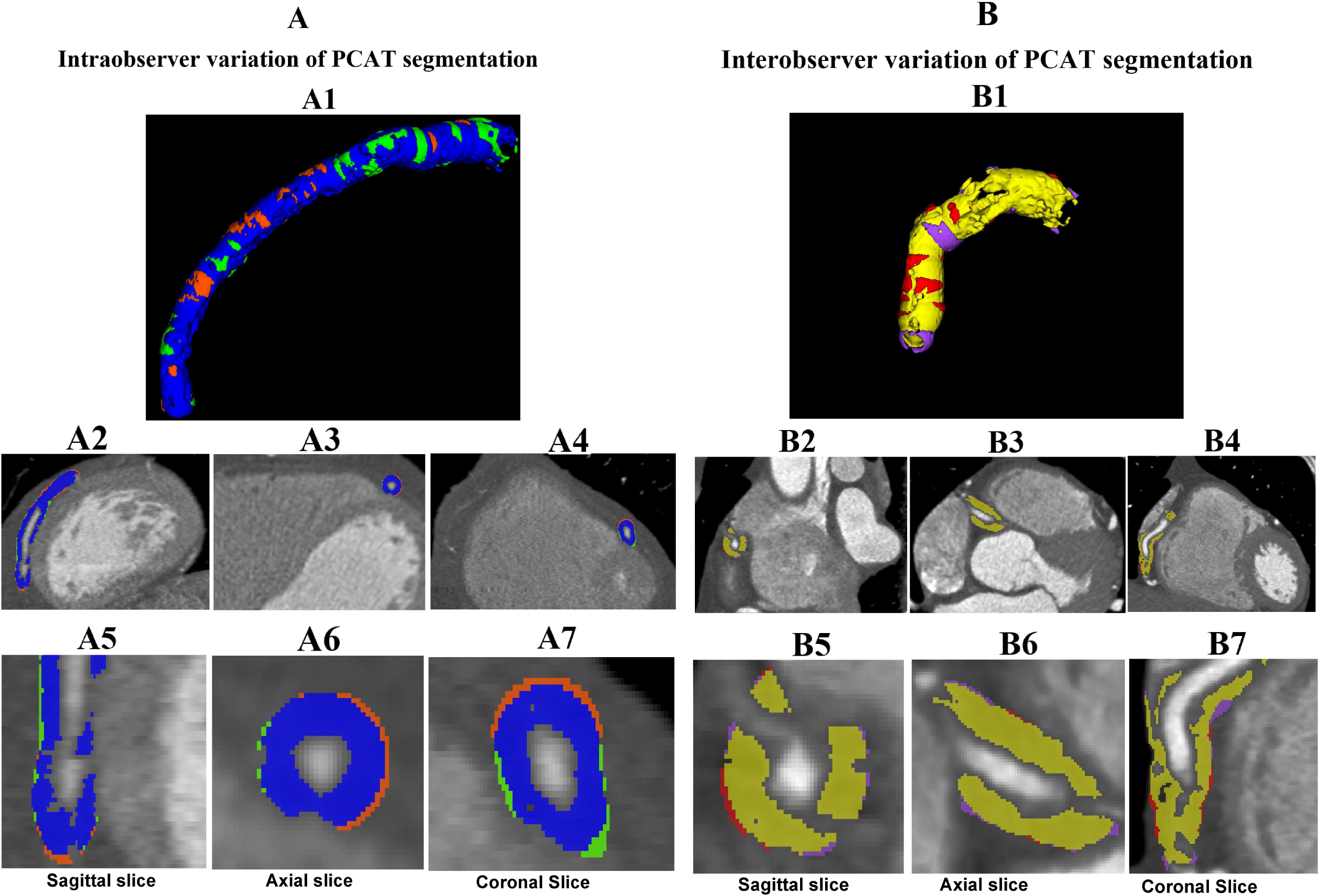
An example of the intraobserver and variation of LAD PCAT. A1: Voxels classified as LAD PCAT during both segmentations are shown in blue, while voxels that were classified as PCAT during first or second segmentation are shown in green and orange, respectively. The resulting intraobserver Dice Similarity Coefficient was 0.89. Panels A2-4: The overlap (blue) and variations (green, orange) of the two repeated segmentations are displayed in sagittal, axial, and coronal planes, respectively. Panels A5-7 are magnified visuals that correspond to panels A2-4. **Figure 2B:** An example of the interobserver variation of RCA PCAT. B1: The interobserver 3D cylinder segmentation of RCA PCAT with interpreter invariant voxels shown in yellow, and interpreter-specific regions shown in red (Interpreter 1) and purple (Interpreter 2). The resulting interobserver Dice Similarity Coefficient was 0.90. Panels B2-4: the overlap (yellow) and the variation (red, purple) are displayed in sagittal, axial, and coronal planes, respectively. Panels B5-7 are magnified visuals that correspond to panels B2-4.

Paired t-tests, comparing the intraobserver PCATrds did not detect any radiodensity differences in any of the CAs that were segmented (all *p*s >0.05) (Table 1A). The intraobserver CoV ranged from 1.3% to 2.7%, with ICCs ranging from 0.98 to 0.99 in the PCATrd of 4 epicardial CAs. The Bland-Altman analysis showed insignificant intraobserver radiodensity variations in the overall PCAT and artery-specific PCAT (all *p*s > 0.05; Fig. 3A).

**Figure 3.**
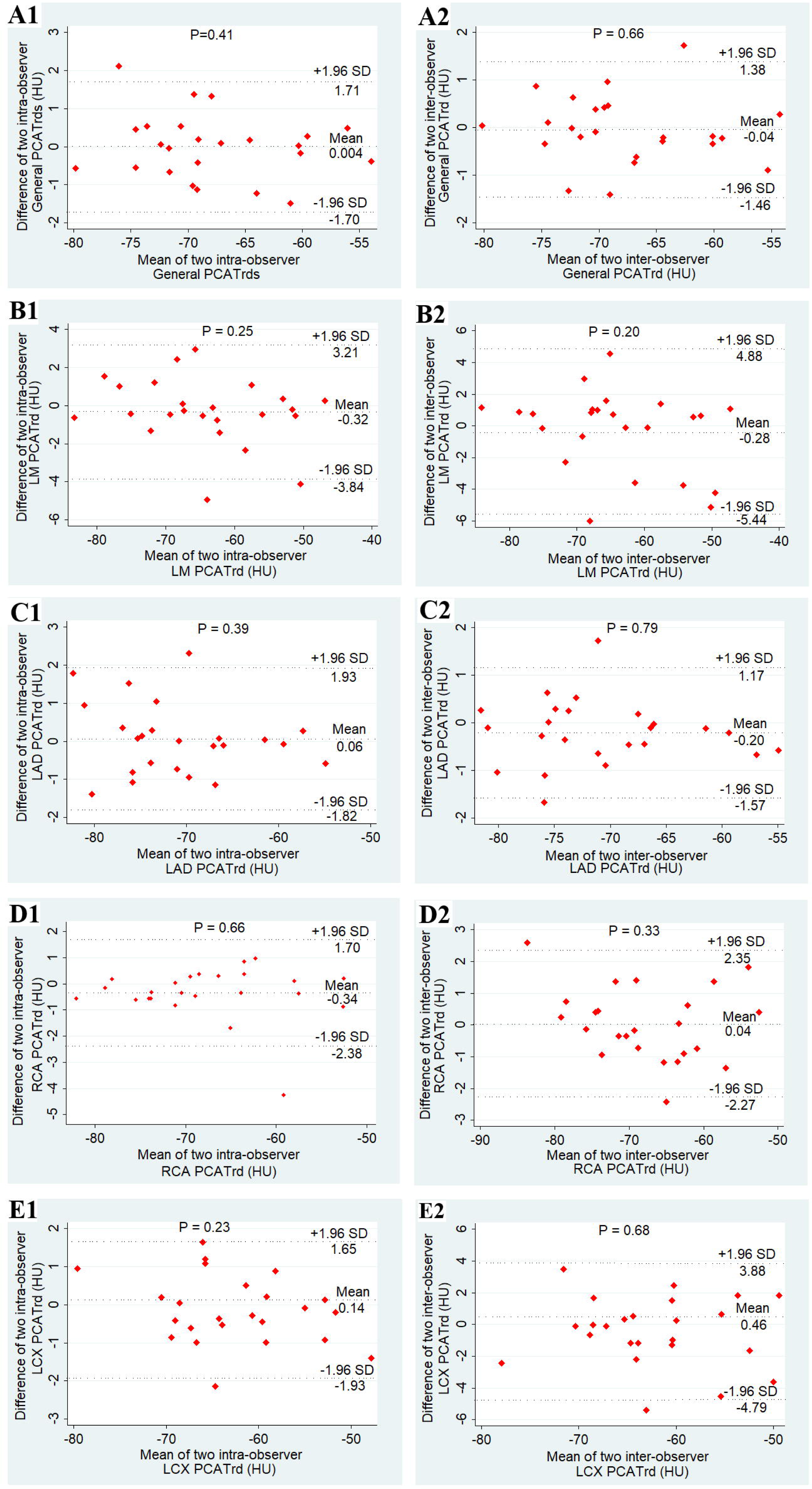
Bland-Altman plot of General PCATrd and PCATrd of 4 major coronary arteries by two intraobserver segmentations (Panel A) and two interobserver segmentations (Panel B) Abbreviations: see Table 1.

### 3.3 Interobserver Reproducibility of PCATrd

The interobserver segmentation overlap (Fig. 2B) was also excellent with DSC 0.84±0.04 for the general PCAT, 0.79±0.04 for the LM PCAT, 0.84±0.04 for the LAD PCAT, 0.85±0.07 for the RCA PCAT, and 0.84±0.04 for the LCX PCAT.

General and artery-specific PCATrds were statistically similar (all *p*s >0.05. The interobserver CoV ranged from 1.0% to 4.0%, with ICCs ranging from 0.96 to 0.99 in the PCATrd of 4 epicardial CAs. The Bland-Altman analysis showed insignificant interobserver variations in PCATrd (all *p*s > 0.05; Fig. 3B).

### 3.4 Reproducibility of PCATrd of the calcified segments

Both the intraobserver DSC and interobserver DSC of the PCAT in calcified segments were excellent, with 0.85±0.07 and 0.83±0.06, respectively. The intraobserver CoV was 1.5%, and ICC was 0.99; The interobserver CoV was 1.3%, and ICC was 0.99 (Table. 2).

**Table 2.** Intraobserver and interobserver measures of PCAT segments.

| **2A. Intraobserver measures (N=25)** |  |  |  |  |  |  |
| --- | --- | --- | --- | --- | --- | --- |
|  | Segmentation overlap | PCATrd (HU) |  |  |  |  |
|  | Dice Similarity Coefficient | Interpreter 1 (Baseline) | Interpreter 1 (3 month later) | P-value | CoV | ICC |
| PCAT | 0.85±0.07 | -68.0±6.1 | -68.1±6.1 | 0.57 | 1.5% | 0.992(0.985, 0.996) |
| **2B. Interobserver measures (N=25)** |  |  |  |  |  |  |
|  | Segmentation overlap | PCATrd (HU) |  |  |  |  |
|  | Dice Similarity Coefficient | Interpreter 1 (Baseline) | Interpreter 2 (Baseline) | P-value | CoV | ICC |
| PCAT | 0.83±0.06 | -68.1±6.1 | -68.0±6.2 | 0.52 | 1.3% | 0.994 (0.988, 0.997) |

Abbreviations: see Table 1.

## 4. Discussion

This study showed that our semi-automated 3D method for segmenting the pericoronary EAT radiodensity (PCATrd) was both feasible and highly reproducible.

We think that this novel technique for quantifying the PCATrd will be valuable for future studies of the relationship between EAT and atherosclerotic disease. Techniques that rely on random sampling of EAT are prone to sampling error because of regional variability in EAT radiodensity [10].Our technique avoids such sampling error because it analyzes the PCATrd for the entire length of main coronary arteries [6; 10]. Our protocol was highly reproducible with excellent intraobserver and interobserver tracing overlap in the major length of CAs. We did not find that the presence of coronary calcification undermined the intraobserver and interobserver reliability. The Dice Similarity Coefficients exceeded our 0.7 cut-offs, the lower boundary for good image overlap, in all reliability assessments, and exceeded 0.8 thresholds for excellent image overlap in all but one reliability test. The CoV, ICC, and Bland-Altman results of the PCATrd were also favorable, further demonstrating that the PCATrd, derived from the proposed 3D segmentation technique, was highly reproducible.

Our protocol represents a highly feasible methodology. The software (i.e., ITK-SNAP) that we used to perform our segmentations is publicly available at no cost and is straightforward to use. It provides synchronized cursor for 3D navigation and allows manual segmentation in three orthogonal planes simultaneously [16]; our research team provides detailed pipeline analysis with step by step instruction allowing easy replication of our technique. The MATLAB code for identifying the EAT via user-specified radiodensity thresholds is provided in the supplementary materials and the threshold range can be easily adjusted to suit project-specific needs. Key considerations of our methodology for PCATrd warrant additional discussion. We used contrast-enhanced CT datasets due to their high signal to noise to delineate CAs. The radiodensity threshold was also used to provide good differentiation of EAT tissue with high edge sharpness between the CAs and EAT. The radiodensity thresholds (i.e., -190,-3 Hounsfield Unit) used in the current study was previously established for quantifying EAT from contrast-enhanced scans, as the conventional radiodensity (i.e., -190,-30 Hounsfield Unit) thresholds significantly underestimate EAT volumes on contrast CT, in comparison to non-contrast CT [15]. However, none of our segmentation rules and landmarks were based on a particular radiodensity threshold. Different EAT thresholding will have minimal, if any effect, on segmentation reliability. We also ensured that our CT dataset had a uniform CT tube voltage and used a consistent contrast agent concentration to minimize their influences on EAT radiodensity[19; 20]. During segmentation, the starting and ending points for continuous tracing needed to be well defined using standardized nomenclature [9]; the orthogonal radius of the PCAT segmentation also needs to be standardized [7].

There are some limitations to this study. Firstly, we did not measure the PCATrd of CA branches, only the four main CAs. However, the authors feel that the additional information gained from analyzing branches is outweighed by the time needed to perform this additional quantification. Second, this semi-automatic segmentation method is relatively time-consuming; however, it is still comparable to other studies [6].

## Future directions

A fully automated segmentation of PCATrd is needed to enable applications for large cohorts. The PCATrd should be correlated with the severity of coronary artery disease and ultimately its association with adverse outcomes should be determined. The PCATrd could also be analyzed at a coronary segment level using IVUS and OCT to study associations of local PCATrd changes with coronary plaques morphology, while longitudinal studies of the PCATrd could be informative of PCATrd changes over time. We also recommend that our 3D segmentation protocol be adapted for non-contrast cardiovascular CT in future studies.

## Conclusion

Our novel 3D segmentation technique to characterized pericoronary EAT radiodensity is feasible, comprehensive, and reproducible. With the open source of the software, plus provided the detailed image analysis pipeline, this segmentation method will be easily and reliably replicated, and have will broad applicability.

## Data Availability

The data can be available upon reseasonal requests.

## Abbreviations

(PCATrd): pericoronary epicardial adipose tissue radiodensity
(EAT): epicardial adipose tissue
(3D): three-dimensional
(DSCs): Dice Similarity Coefficients
(ICC): intraclass coefficient
(CA): coronary artery
(CCTA): coronary computed tomography angiography
(LM): left main
(LAD): left anterior descending
(LCX): left circumflex
(RCA): arteries right coronary artery

## Compliance with ethical standards

Guarantor The scientific guarantor of this publication is Craig Butler.

### Statistics and biometry

There were no complex statistical methods necessary for this paper.

### Informed consent

Written informed consent was obtained from all subjects (patients) in this study.

### Ethical approval

Institutional Review Board approval was obtained.

### Methodology

- Retrospective
- performed at one institution

---

Supplemental material 1. The pipeline of the PCAT segmentation

**Supplemental Figure 1.**
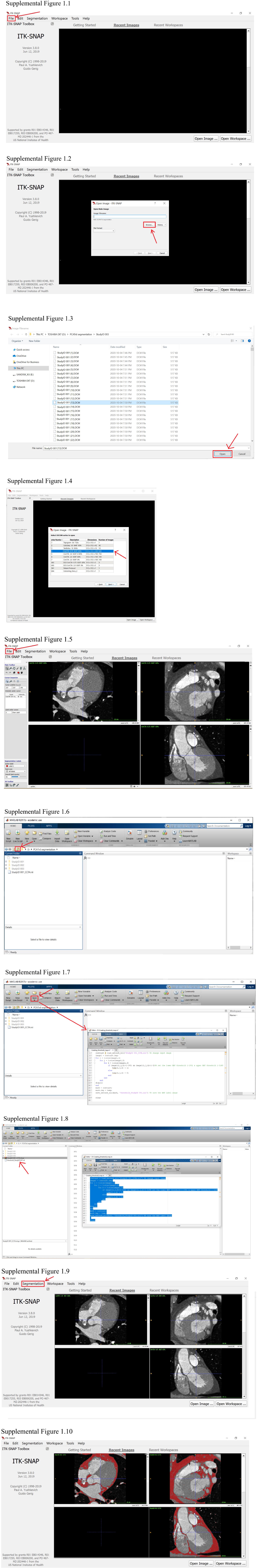
The pipeline of the PCAT segmentation: the pre-segmentation preparation.

**Supplemental Figure 2.**
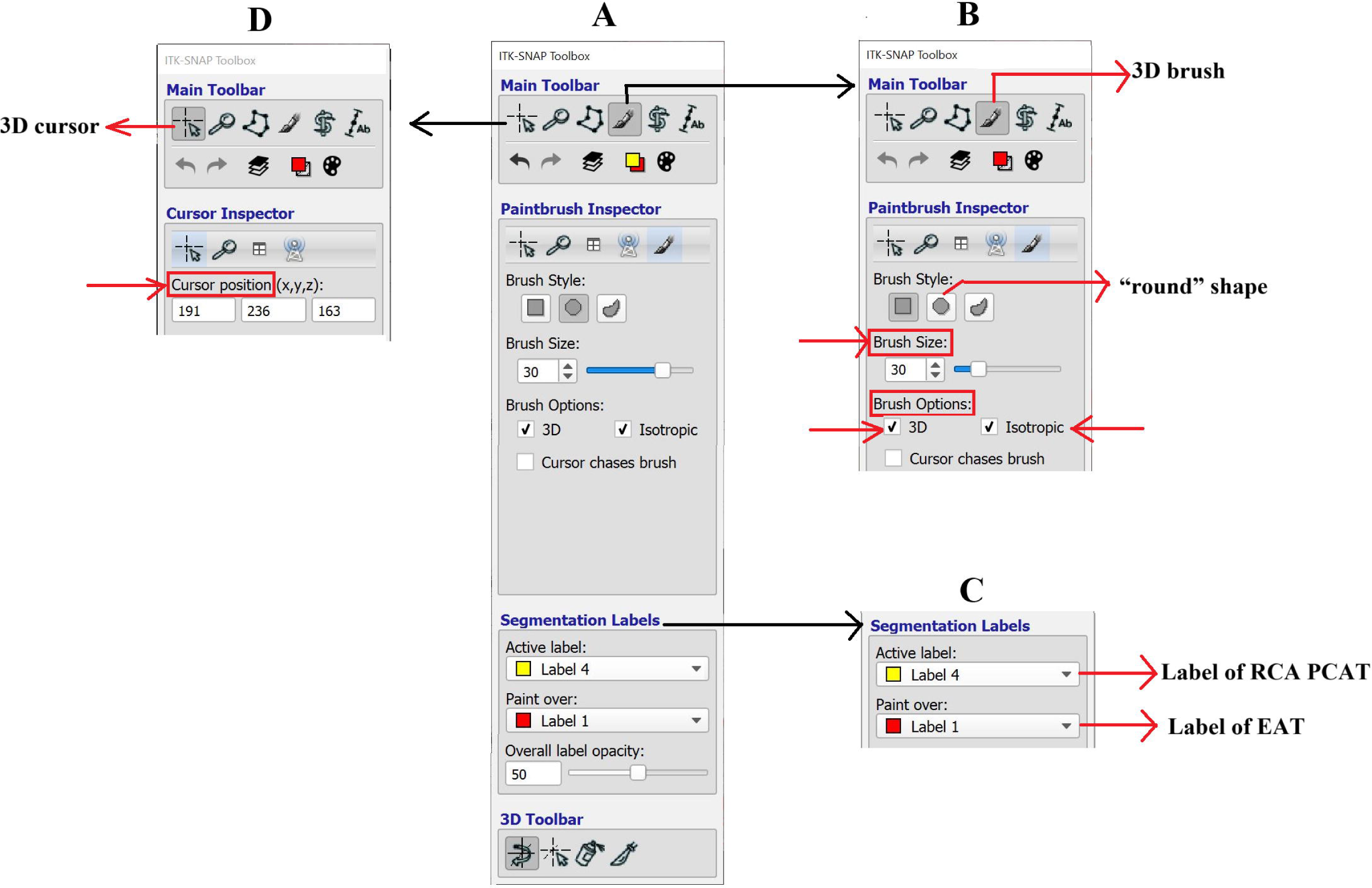
Introduction of segementation tools in ITK-SNAP software.

## Supplemental material 1: the pipeline of the PCAT segmentation

Step 1. Save the CCTA image series using NIFTI format in ITK-SNAP software.

### 1) Install the ITK-SNAP software

Navigate to the website: “http://www.itksnap.org/pmwiki/pmwiki.php?n=Downloads.SNAP3” and download ITK-SNAP software version appropriate for your operating system and install it.

### 2) Upload the CT dataset

The CT datasets can be uploaded to ITK-SNAP in a variety of formats including DICOM, NIFTI, MetaImage or NRRD.[21]

To open the dataset in ITK-SNAP, click the “File” toolbar---“Open main image” (See supplemental Figure 1.1) and browse (See supplemental Figure 1.2) to the folder containing the saved CT dataset to be analysed (e.g., *“StudyID 001”*). Select the target dataset, click “Open” (See supplemental Fig 1.3). All image series contained in the CT dataset could be uploaded and displayed in ITK-SNAP (See supplemental Figure 1.4).

### c) Save the image of CCTA series as NIFTI file

Select the desired CCTA image series from the available series (e.g., the series *“CorCTA 0.75 B25f 65”* in current study), and click “Next” (See supplemental Figure 1.4). The selected series was displayed as a multiplanar reformat (i.e. axial, coronal and sagittal orientations) (See supplemental Figure 1.5). Finally, click “File” ---“Save image” ---“Main image” in the tool bar (See supplemental Figure 1.5) to save the CCTA series and specify a file name (i.e., *StudyID 001_CCTA.nii*) and a file location using a NIFTI format file.

Step 2. Create the Hounsfield Unit defined threshold map of epicardial adipose tissue in MATLAB software.

a. In MATLAB software, set the directory to the folder where “*StudyID 001_CCTA.nii”* was saved by clicking the “browse the folder” icon (See supplemental Figure 1.6).
b. Open the MATLAB code for creating the threshold map by clicking “Open” in the toolbar and select the MATLAB file (e.g., file name “*Creating_threshold_map.m*”) to create radiodensity-based EAT threshold map (See supplemental Figure 1.7). The MATLAB code “*Creating_threshold_map.m*” below has been written according to NifTi read/write functions (https://www.mathworks.com/matlabcentral/fileexchange/8797-tools-for-nifti-and-analyze-image). Only the text **highlighted in purple** should be customized for each dataset. The “-190” and “-3” were the lower limit and the upper limit of the attenuation threshold for EAT, respectively. These can be customized for individual need and preferences.

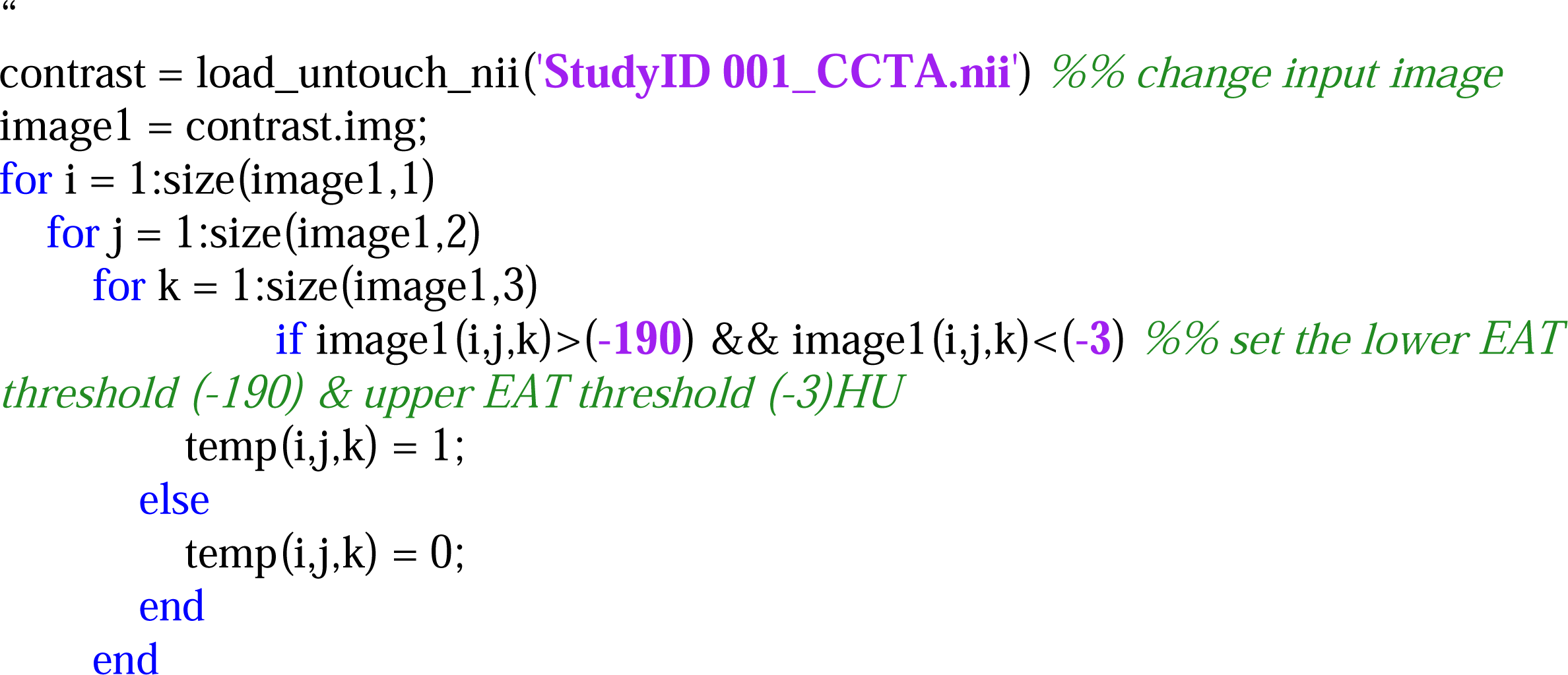
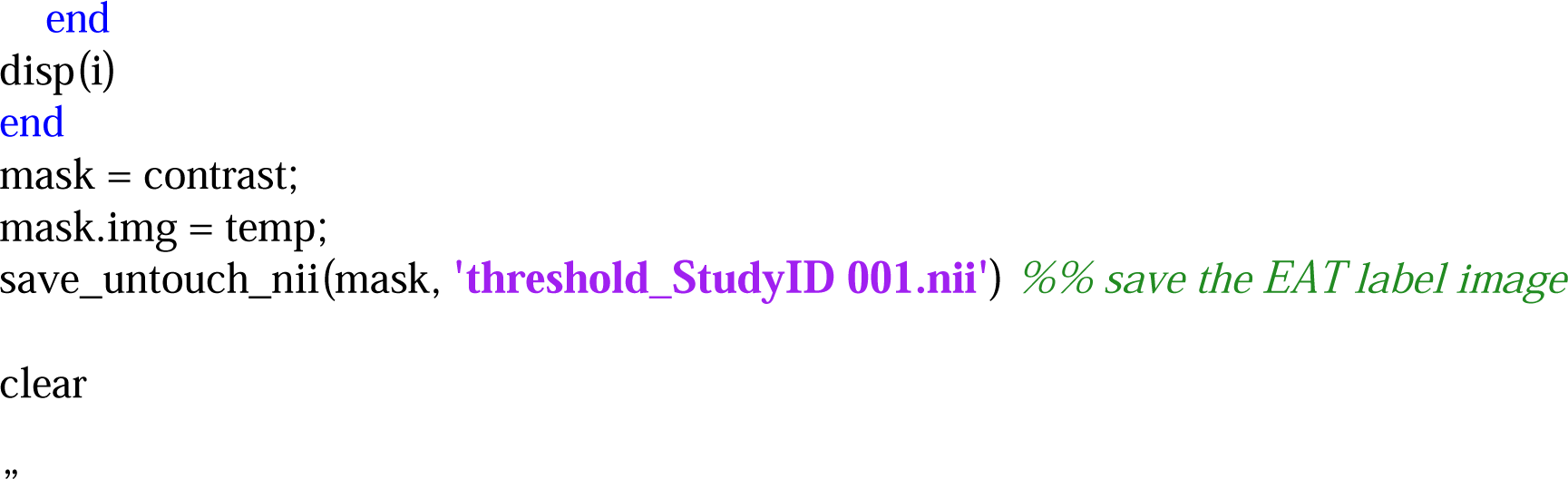
c. To run the MATAB code, select the code, right click the selected code, and select “Evaluate selection” (See supplemental Figure 1.8). Then the threshold map *’threshold_StudyID 001.nii’* for the input image ‘*StudyID 001_CCTA.nii’* was created (See supplemental Figure 1.8).

Step 3. Apply threshold map to CCTA image in ITK-SNAP software

a. Upload the CCTA image (*’StudyID 001_CCTA.nii’*) in ITK-SNAP software by clicking “File” in the toolbar ---“Open main image”---“Browse” (See supplemental Figure 1.1 & 1.2) and select the targeted CCTA image (‘*StudyID 001_CCTA.nii’*); The ‘*StudyID 001_CCTA.nii’* was three-dimensionally displayed (See supplemental Figure 1.5).
b. Upload the threshold map (*’threshold_StudyID 001.nii’*) in the ITK-SNAP software by clicking “Segmentation” in the toolbar---“Open segmentation”---“Browse” and select the *’threshold_StudyID 001.nii’* (See supplemental Figure 1.9). Then EAT threshold map *’threshold_StudyID 001.nii’* was applied to the CCTA image *’StudyID 001_CCTA.nii’* (See supplemental Figure 1.10). The adipose tissue was highlighted in red on the image.

Step 4. Segment the PCAT in ITK-SANP software.

With the CCTA series and EAT threshold map now displayed in the ITK-SNAP multiplanar viewer, segmentation can begin. We defined PCAT as the EAT adjacent to the CAs, in a three-dimensional cylinder with a radial distance from the outer coronary vascular wall equal to the coronary diameter [6; 7] of the corresponding coronary segment. PCAT was manually segmented in three orthogonal planes simultaneously (Figure 1A1-A3) in paintbrush mode using a spherical brush, with ‘round,’ ‘3D,’ and ‘isotropic’ settings (Supplemental Fig 2B) with an appropriate brush size. Tracing (i.e., the “Active label” in Supplemental Fig 2C) was performed on top of the previously identified EAT tissue label (i.e., the “Paint over” label in Supplemental Fig 2C). The 3D brush’s cursor (Supplemental Fig 2D) was placed at the center of each CA segment (e.g., RCA in Fig 1) in three orthogonal planes simultaneously (Fig 1. A1-A3). By having a left click, the PCAT was simultaneously segmented in three orthogonal planes (Fig 1. A1-A3). Then relocate the 3D cursor along the CA with 2-3 mm intervals between two consecutive tracings (Fig 1B1-B2) and ensure 3D cursor at the center of CA segment in three orthogonal planes simultaneously. Adjust the brush size if necessary and repeat the segmentation. The PCAT of the left main (LM) was segmented from the ostium of the LM to the bifurcation origins of the left anterior descending (LAD) and left circumflex (LCX) arteries (SCCT Segment 5). The segmenting path of the LAD PCAT began at the LM bifurcation, traveled in the anterior interventricular groove, and ended at the LV apical terminal (SCCT Segment 6-8). The LCX PCAT was segmented in the atrioventricular groove from its origin to the end of the vessel or to the origin of the left posterior descending artery (SCCT Segment 11 and Segment 13). Lastly, PCAT of the right CA (RCA) was segmented from the ostium of the RCA to the origin of the posterior descending artery (SCCT Segment 1-3) [8; 9] (Fig. 1C1-C3). The segmentation stopped when at the aforementioned termination points or if the diameter of a given CA narrowed to less than 2 mm [22]. A 3D visualization of segmented PCAT from one of our participants is shown in Fig. 1D. The PCATrd was computed by averaging radiodensity values across all voxels belonging to a given PCAT label. Finally, we computed general PCATrd by collapsing all 4 PCAT segments (i.e., LM, LAD, LCX, and RCA) into a single label Fig. 1D1-D3.

## References

1 Talman AH, Psaltis PJ, Cameron JD, Meredith IT, Seneviratne SK, Wong DT (2014) Epicardial adipose tissue: far more than a fat depot. Cardiovasc Diagn Ther 4:416–429

2 Zobel EH, Christensen RH, Winther SA et al (2020) Relation of cardiac adipose tissue to coronary calcification and myocardial microvascular function in type 1 and type 2 diabetes. Cardiovasc Diabetol 19:16

3 Goeller M, Achenbach S, Marwan M et al (2018) Epicardial adipose tissue density and volume are related to subclinical atherosclerosis, inflammation and major adverse cardiac events in asymptomatic subjects. J Cardiovasc Comput Tomogr 12:67–73

4 Franssens BT, Nathoe HM, Leiner T, van der Graaf Y, Visseren FL, group Ss (2017) Relation between cardiovascular disease risk factors and epicardial adipose tissue density on cardiac computed tomography in patients at high risk of cardiovascular events. Eur J Prev Cardiol 24:660–670

5 Abazid RM, Smettei OA, Kattea MO et al (2017) Relation Between Epicardial Fat and Subclinical Atherosclerosis in Asymptomatic Individuals. J Thorac Imaging 32:378–382

6 Antonopoulos AS, Sanna F, Sabharwal N et al (2017) Detecting human coronary inflammation by imaging perivascular fat. Sci Transl Med 9

7 Oikonomou EK, Marwan M, Desai MY et al (2018) Non-invasive detection of coronary inflammation using computed tomography and prediction of residual cardiovascular risk (the CRISP CT study): a post-hoc analysis of prospective outcome data. Lancet 392:929–939

8 Raff GL, Abidov A, Achenbach S et al (2009) SCCT guidelines for the interpretation and reporting of coronary computed tomographic angiography. J Cardiovasc Comput Tomogr 3:122–136

9 Leipsic J, Abbara S, Achenbach S et al (2014) SCCT guidelines for the interpretation and reporting of coronary CT angiography: a report of the Society of Cardiovascular Computed Tomography Guidelines Committee. J Cardiovasc Comput Tomogr 8:342–358

10 Hell MM, Achenbach S, Schuhbaeck A, Klinghammer L, May MS, Marwan M (2016) CT-based analysis of pericoronary adipose tissue density: Relation to cardiovascular risk factors and epicardial adipose tissue volume. J Cardiovasc Comput Tomogr 10:52–60

11 Walter SD, Eliasziw M, Donner A (1998) Sample size and optimal designs for reliability studies. Stat Med 17:101–110

12 Bonett DG (2002) Sample size requirements for estimating intraclass correlations with desired precision. Stat Med 21:1331–1335

13 Koo TK, Li MY (2016) A Guideline of Selecting and Reporting Intraclass Correlation Coefficients for Reliability Research. J Chiropr Med 15:155–163

14 Sheth T, Butler C, Chow B et al (2012) The coronary CT angiography vision protocol: a prospective observational imaging cohort study in patients undergoing non-cardiac surgery. BMJ Open 2

15 Xu L, Xu Y, Coulden R et al (2018) Comparison of epicardial adipose tissue radiodensity threshold between contrast and non-contrast enhanced computed tomography scans: A cohort study of derivation and validation. Atherosclerosis 275:74–79

16 Yushkevich PA, Piven J, Hazlett HC et al (2006) User-guided 3D active contour segmentation of anatomical structures: significantly improved efficiency and reliability. Neuroimage 31:1116–1128

17 Zou KH, Warfield SK, Bharatha A et al (2004) Statistical validation of image segmentation quality based on a spatial overlap index. Acad Radiol 11:178–189

18 Zijdenbos AP, Dawant BM, Margolin RA, Palmer AC (1994) Morphometric analysis of white matter lesions in MR images: method and validation. IEEE Trans Med Imaging 13:716–724

19 Coppini G, Favilla R, Marraccini P, Moroni D, Pieri G (2010) Quantification of Epicardial Fat by Cardiac CT Imaging. Open Med Inform J 4:126–135

20 McCollough CH, Leng S, Yu L, Fletcher JG (2015) Dual- and Multi-Energy CT: Principles, Technical Approaches, and Clinical Applications. Radiology 276:637–653

21 Yushkevich PA, Pashchinskiy A, Oguz I et al (2019) User-Guided Segmentation of Multi-modality Medical Imaging Datasets with ITK-SNAP. Neuroinformatics 17:83–102

22 Sundaram B, Patel S, Agarwal P, Kazerooni EA (2009) Anatomy and terminology for the interpretation and reporting of cardiac MDCT: part 2, CT angiography, cardiac function assessment, and noncoronary and extracardiac findings. AJR Am J Roentgenol 192:584–598

